# The development of a guidance document for interprofessional collaborative advance care planning in dementia care

**DOI:** 10.1101/2023.01.25.23285020

**Authors:** C. Khemai, D.J.A. Janssen, J. Van Dongen, J.M.A. Biesmans, S.R. Bolt, J.M.G.A. Schols, J.M.M. Meijers

## Abstract

Advance care planning (ACP) is an essential part of palliative dementia care and should embrace a collaborative approach involving persons with dementia, family members and various healthcare professionals (HCPs). This study aimed to develop a guidance document for IPC in ACP in dementia care for HCPs from nursing homes, using a mixed-method design and taking an appreciative inquiry approach comprising semi-structured focus groups and interviews with HCPs. The study participants (N=18) included certified nurses, registered nurses, a quality nurse, nurse specialists, team leaders, physicians specialized in old age medicine, psychologists, and a teacher/researcher (mean age 43.8 years; 79% female). The data analysis indicated six elements to include in the guidance document. The first three elements concerned common awareness of collaborative ACP in dementia care, role clarification for various professions, and actively working towards shared goals in collaborative ACP in dementia care. The last three themes focused on the dynamic processes in collaborative ACP in dementia care: interprofessional communication, coordination, monitoring and evaluation. This study provides a guidance document that HCPs could use in practice to identify the elements they can address to improve collaborative ACP in dementia care.

## Introduction

Dementia is a neurodegenerative disease with disorders in at least two cognitive domains, which leads to functional impediments to daily life^1^. The progressive and incurable character of dementia^2^ asks for timely advance care planning (ACP), which is defined as a process *‘to enable individuals to define goals and preferences for future medical treatment and care, to discuss these goals and preferences with family and healthcare professionals (HCPs), and to record and review these preferences if appropriate’* ^3^. Proxy ACP is used for persons with advanced dementia who have lost their decision-making capacity^4^. In proxy ACP, the proxy decision-maker (often a family member) is involved in the conversations and has the ultimate legal responsibility^4^. Timely ACP involving the person with dementia is needed before they lose their decision-making capacity^5^ in order to prepare family members for this challenging task^6^, and ensure that they can make end-of-life decisions in line with the preferences of their relative with dementia^7^.

ACP is known to decrease burdensome care movements and medical treatments^8-10^, maintain autonomy^11, 12^, improve overall end-of-life care^13^, and improve the quality of palliative care (PC)^14^ for persons with dementia. Despite the importance of initiating ACP at an early stage due to cognitive decline^15, 16^ and preferably when still living at home^17^, ACP is rarely initiated among persons with dementia in the home care setting^17, 18^. As 92% of persons with dementia die in a nursing home (NH)^19^, with a median survival rate of two and a half years after the move^20^, it is necessary to conduct or/and evaluate ACP in NHs ideally prior to^21^, or soon after the move^22^. Nevertheless, ACP (involving family members) is not enacted for every Dutch resident with dementia directly after the move to the NH^23, 24^.

ACP is considered as a shared responsibility of persons with dementia^5^, family members^25^ and the interprofessioneal team^26, 27^. For example, physicians^28, 29^, nurses^28, 30-32^, psychologists^33^, social workers^28, 34^, and priests^28, 35^. To date, a few guidances or practical tools on how to conduct ACP conversations in dementia care have been documented^36-38^. However, there is no guidance for IPC in ACP for persons with dementia exists. This study thus aimed to develop such guidance for HCPs, by describing the essential elements for IPC regarding ACP in dementia care.

## Methods

### Design and setting

This study is part of the larger Desired Dementia Care Towards End of Life (DEDICATED) research project, which aims to improve quality of PC for persons with dementia living at home or in a NH, and during the move from home to the NH^39^. The current study describes the development of the *DEDICATED collaborative approach for advance care planning in dementia care* guidance document. The overall development process consisted of five steps (Fig 1 and online Supplement I). The present article describes the final step of the development process. A mixed-method design and appreciative inquiry approach were used. The Consolidated criteria for Reporting Qualitative research (COREQ)^40^ was followed (Supplement II). Appreciative inquiry compromises four phases: the Discovery phase, Dream phase, Design phase, and Destiny phase^41^.

**Fig 1.**
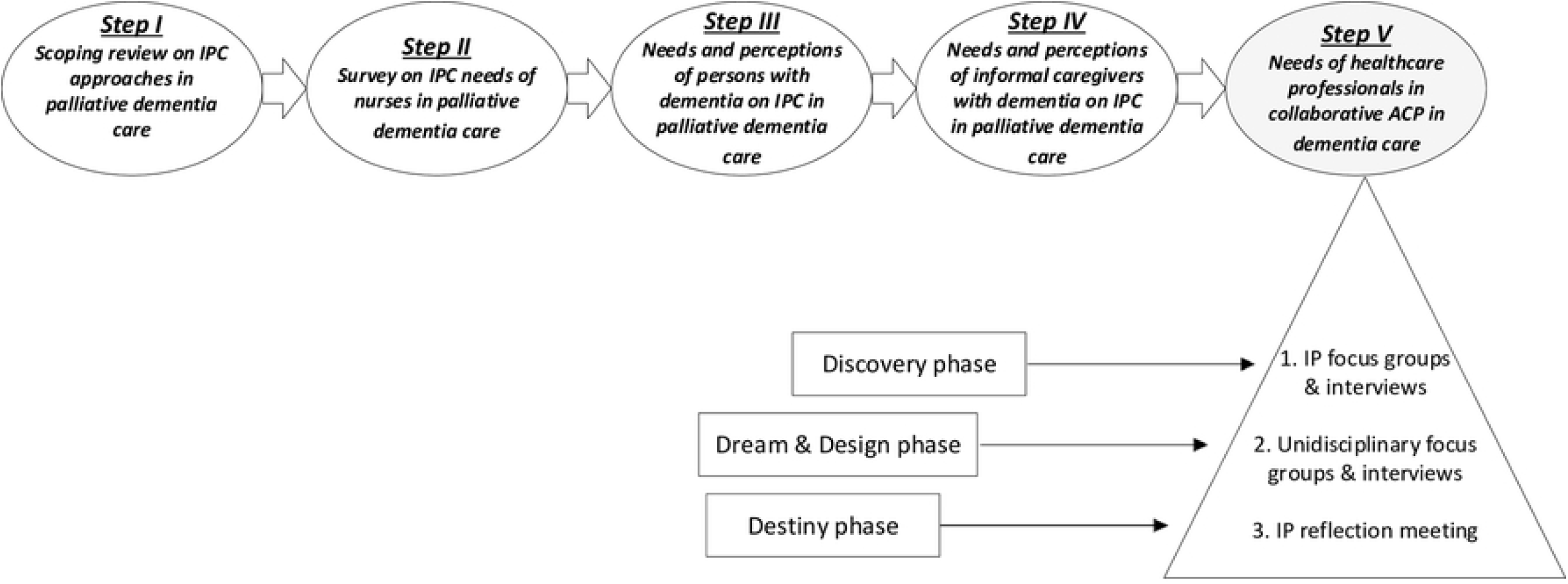
Development process of the guidance document for DEDICATED collaborative advance care planning in dementia care. Abbreviations: IPC = Interprofessional collaboration; ACP = Advance care planning; IP = interprofessional.

First, we undertook the Discovery and Dream phases of the appreciative inquiry. During the interprofessional focus groups and interviews with nurses, physicians, team leaders and psychologists from NHs, we focussed on exploring the current state of and their wishes and needs in collaborative ACP in dementia care (the Discovery phase of the appreciative inquiry approach). Next, we conducted unidisciplinary focus groups and interviews to discuss, elaborate and refine the initial results (the Dream and Design phases of the appreciative inquiry approach). During these unidisciplinary focus groups and interviews, we also clarified how the diverse roles of the HCPs complemented each other in ACP. We asked for further explanations about the identified themes and for possible solutions to challenges in collaborative advance care planning in dementia. Finally, we discussed the elements of the *DEDICATED collaborative approach for advance care planning* guidance document in an interprofessional reflection meeting in which participants provided their feedback on the selected elements (Destiny phase of the appreciative inquiry approach). After development of the guidance document, we performed member checks in which we asked the participants to check for accuracy and whether the information provided resonated with the input they provided^42^.

### Sampling, data collection and data analysis

We used different non-probability purposive and snowballing sampling techniques to recruit participants for Step V^43^. During every interview and focus group, two female DEDICATED-team members were involved. Detailed information about data collection is provided in Supplement III. All participants provided verbal and written informed consent. The semi-structured interview guide used during the development process was based on literature describing crucial PC elements in dementia care^44, 45^, aspects of IPC^46-51^, IPC regarding ACP^8, 52-55^, existing assessment tools and questionnaires about IPC^56, 57^, and appreciative inquiry phases (Supplement IV). No field notes were made.

Information was audio recorded and transcribed in clean verbatim style. C.K. undertook inductive content analysis by identifying meaning units and condensing them from the transcripts, and developing codes, categories and themes^58^. These findings were discussed with J.M.M. and the participants from the interviews and focus groups to ensure the internal reliability of the data^59^. The coding tree is provided in Supplement V. We reached code saturation at the eight transcript (first unidisciplinary focus group) and meaning saturation at the tenth transcript (third unidisciplinary duo-interview). Transcript were not returned to the participants for comments or corrections.

## Results

Five of the twenty-three candidates declined to participate due to high workload. Eighteen participants contributed to Step V of the development process of the guidance document on collaborative ACP in dementia care: two certified nurses, three registered nurses, one quality nurse, two nurse specialists, three team leaders, three elderly care physicians, three psychologists, and one teacher/researcher (Supplement VI). The participants had a mean age of 43.8 years (mean 42.5 years; range 55.0 years) and were predominantly women (79%). Approximately 40% of the participants had at least ten years of work experience in their current profession. Each interprofessional focus group or interview lasted approximately one hour and 11 minutes. The mean interview time for unidisciplinary focus groups or interviews was one hour and 48 minutes. The reflection meeting lasted for two hours and five minutes. The identified themes are depicted in Fig 2. The key quotes of the participants are displayed in Table 1.

**Table 1.** Key quotes of the participants.

| Theme | Key quote(s) |
| --- | --- |
| *Theme 1. The meaning of advance care planning* | *“Advance care planning is actually what I ask persons with dementia and their families: ‘What should I know from you in order to provide preferred care, not just right now at this moment, but also later on if you are not doing so well’”. (Nurse Specialist)* |
| *Theme 2. The timing and continuity of advance care planning* | *“These advance care planning conversations should be conducted in an earlier phase. Currently, these conversations are conducted when something happens. That is too late”. (Certified Nurse Assistant)*<br>*“Advance care planning conversations should be conducted as soon as possible, but should also be customised to the person with dementia and their family members, and be conducted on a regular basis. Otherwise, there is a danger that people think it is just a task that should be checked off and then you are done with it.” (Physician specialized in old age medicine)* |
| Theme 3. Lacking information from the home care setting | *“Advance care planning information is often not present in patient files and transferred during the NH admission. During the file transfer, information regarding general healthcare domains such as physical, participation, mental wellbeing etc. is often documented, but nothing is mentioned about their personal life, or wishes and needs regarding the end-of-life.” (Psychologist)* |
| Theme 4. Creating role clarity in advance care planning | *“Advance care planning is a subject in which I am not really involved. I also asked my colleagues about this subject, and this is something in which currently the physician plays a major role, but not psychologists. There is also scarce literature about the role of psychologists in ACP. So, I really think that we need to define the role of a psychologist in ACP.” (Psychologist)* |
| Theme 5. Having shared visions | *“We actually never talked in our team about our mission in advance care planning”. (Team leader)* |
| Theme 6. Sharing perspectives & making shared-decisions | *“In the end, it is the physician or/and nurse specialist who officially makes the final decision and it depends on how approachable the physician is about receiving your input and how assertive you are to about making a shared decision. With most physicians, I make shared-decisions; however, in some rare cases, I did encounter authoritative physicians, and then I noticed that as a nurse or psychologist, it becomes difficult to say anything.” (Psychologist)*<br>*“It is important to discuss care aspects in an early phase with persons with dementia and talk about the things they want, but in many cases when the situation changes, you still have to conduct conversations at that moment with* |
|  | *the client and family member in order to make a choice, which is difficult to secure. Eventually, the physician has the final say. ” (Certified Nurse Assistant)* |
| *Theme 7. From advance care planning information to actions* | *“The information that is discussed during the meetings (i.e. multidisciplinary team, resident, physician-nurse and psychologist-nurse meetings) is included in the reports, but you do not have the time to verbally explain the reasoning behind the agreements that are made to all colleagues.” (Registered Nurse)* |
| *Theme 8. Having a facilitating organisation* | *“I plead for regular short verbal information transfers and resident meetings. In practice, in my opinion, we write everything, but let’s be honest, if you read the reports of 20 residents, you have already forgotten about the first five when you to start with your shift. When someone tells you, what is important and what not to provide suitable care, than it is easy to memorize. But now these important verbal information transfer moments have disappeared from our organization.”*<br>*(Nurse Specialist)* |
| *Theme 9. Evaluating the interprofessional collaboration process and end-of-life care* | *“There was a case in which a resident had an euthanasia statement, but was declared incompetent. We all agreed not to execute any life-prolonging treatments. However, during an acute situation, the physician on duty still decided otherwise and family was fine with it, but this action was not in accordance with our NH policy. This is a good case to evaluate, because there was a shared-decision, but still a different action was executed: how did that happen and how can we learn from this?” (Team leader)* |

**Fig 2.**
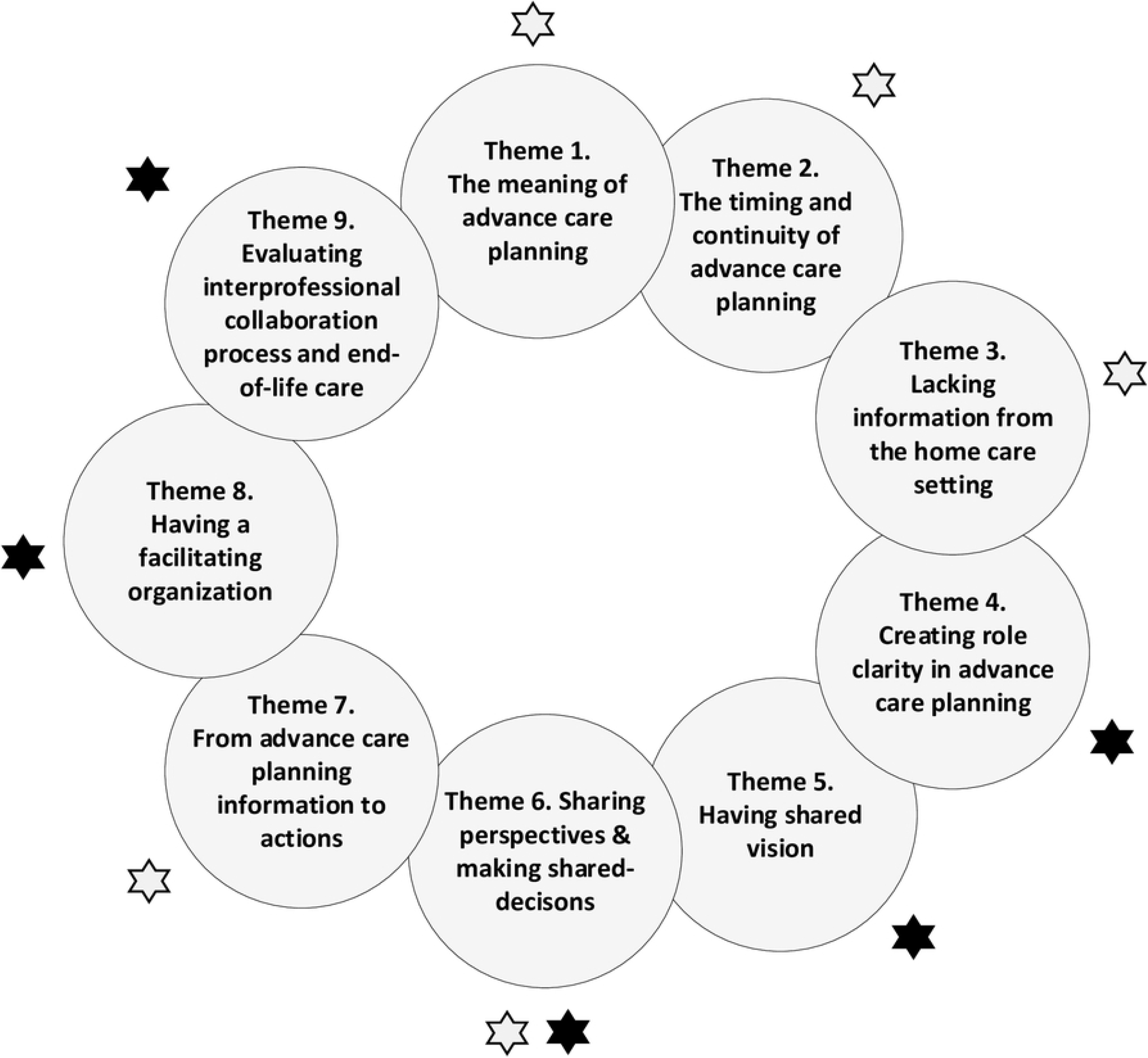
Identified themes that describe the elements for optimal interprofessional collaboration in advance care planning for persons with dementia living in nursing homes. The white asterisks refer to the themes that arose from the interprofessional focus groups and interviews. The black asterisks refer to the themes that arose from the unidisciplinary focus groups and interviews.

### Theme 1. The meaning of ACP

Most nurses and psychologists indicated that they either had not heard about ACP or did not structurally use the term in daily practice. The majority emphasized that many of their colleagues did not realize that dementia is a life-limiting disease and that PC, with ACP as one of the components, is applicable before the end-of-life. They also mentioned the stigma around PC, because many HCPs, persons with dementia and family members often misconceive PC as a synonym of end-of-life care. Others brought up the existence of different terms for ACP, such as proactive care planning, end-of-life planning or future care planning, which can cause confusion. All participants suggested using one term (ACP), and noted that the term ACP does not only focus on medical aspects. The participants described ACP as a necessary process to improve autonomy and person-centred care directly after NH admission and during the end-of-life phase of persons with dementia in NHs. They underlined the importance of communicating realistic expectations about living in a NH, as most residents with dementia had reached an advanced stage at time of admission. Discussing preferences regarding life-sustaining treatments (e.g. resuscitation, hospitalisation and mechanical ventilation), medical palliative treatments (e.g. morphine and palliative sedation), psychosocial aspects (e.g. coping with life and death), and spiritual wishes (e.g. rituals during end-of-life) were reported as part of ACP.

### Theme 2. The timing and continuity of ACP

Participants mentioned the importance of conducting ACP in an early phase, preferably in the home care setting with the person with dementia and their family, due to the increasing cognitive decline of a the person with dementia. The participants said that in most cases ACP did not take place in the home care setting. They usually conducted ACP conversations between two and six weeks after the move. However, they explained that in daily practice these conversations are usually initiated at the sixth week after the moving day and often after common adverse events during dementia care, such as pneumonia. Moreover, ACP conversations in NHs are only conducted with family members, because most persons with dementia were partly or not at all able to participate. Because of lack of timely ACP, the family needs to make acute treatment decisions during common adverse events, which causes distress in family members. According to the participants, ACP conversations should ideally occur prior to the move.

Participants noted that the focus of ACP should be on the conversations themselves, rather than solely making decisions and documenting end-of-life wishes. ACP conversations should be ongoing conversations, as the needs and wishes of persons with dementia and their family change during the care trajectory. Participants also emphasized the importance of identifying the coping styles used by persons with dementia and their families, because this coping style forms the basis for conducting customized ACP conversations, adjusted to their willingness, acceptance, expectations, wishes and needs.

### Theme 3. Lacking information from the home care setting

The participants unambiguously said that ACP information if often lacking in file transfer during the move, which means that to the wishes and needs of persons with dementia remain unknown, as most persons with dementia suffer from a limited ability to express their preferences when they arrive at the NH. Receiving ACP information from the home care setting was valued, because it served as a basis from which to conduct follow-up ACP conversations with persons with dementia and family members. Transferring ACP information also prevents family members having to repeat information and make unprepared end-of-life decisions in acute situations. Many participants explained that either the client counsellor (HCP who facilitates the NH transition) or designated nurse assistant from the NH could also play a role in retrieving information about end-of-life wishes prior to the move.

### Theme 4. Creating role clarity in ACP

Collaborating with other disciplines to integrate diverse perspectives was perceived as valuable, however, most participants explained that that ACP in current practice is mostly limited to nurse specialists, physicians specialized in old age medicine, and spiritual caregivers. All participants mentioned that expertise from other HCPs in NHs, such as client counsellors, spiritual caregivers, activity therapists, music therapists, and other allied healthcare professionals, is necessary. The participants mainly brought up the issue of the existing lack of role clarity within ACP, and the lack of proactively consulting each other.

General as well as discipline-specific roles can be distinguished regarding the role division among nurses, psychologists, physicians, and team leaders. A general role (e.g. a role that all HCPs execute) includes being the HCP who conducts the ACP conversation. Most important nurse-specific roles include acting as first contact person or trust person, getting to know the person (i.e. making life books) and acting as a central coordinator by signalling diverse HCPs to initiate or play a role in ACP. Core tasks mentioned by psychologists included providing psycho-education about ACP and support to family members and communicating with persons with dementia and family members about psychosocial needs and wishes regarding ACP. Physicians specialized in old age medicine and nurse specialists are responsible for the medical care of the client, making decisions about treatments, making referrals to other HCPs, and explaining the ACP policies. Compared to all the above HCPs, team leaders particularly ensure optimal conditions for IPC; monitor IPC; mediate between nurses, and physicians and allied healthcare professionals; coach the care team; and coordinate IPC at a department or team level with respect to ACP.

The participants stated that regardless of the willingness or preparedness of the persons with dementia and/or their family, HCPs should at least introduce ACP and provide information about its importance. All participants revealed a lack of structured collaborative ACP within their care organisations and wished for a clear overview in which all HCPs are assigned a position and task. Most participants agreed with the procedure in which the nurse assistant who acts as first contact person conducts the first ACP conversation in which they for example, introduce the subject of ACP, identifies the proxy decision-maker and discusses the expectations of the persons with dementia and their family. This designated nurse assistant should subsequently conduct the second ACP conversation together with the elderly care physician and discuss medical-related topics such as resuscitation. Afterwards, other HCPs such as psychologists and spiritual caregivers could carry out ACP conversations, and all involved HCPs should keep each other informed about the ACP conversations conducted, and any agreements made.

### Theme 5. Having shared visions

From all the participants, team leaders expressed the need to develop a team vision and mission in order to make sure everyone is on the same page and works in a similar way towards a shared outcome in ACP. Team leaders explained that they may not be directly involved in cases, but they have a important role in standing strong for the team vision and mission, and have a clear role in supporting HCPs in providing PC. During the focus groups and interviews, the participants developed a vision and mission, core values, objectives and specific action points to work towards a shared outcome for IPC in ACP (Supplement VII). The participants explained that according to their team vision for ACP in dementia care, quality of life and the end-of-life of persons with dementia should be treated as having equal importance. They noted their mission to achieve the shared involvement of persons with dementia, family members and diverse HCPs for the optimal wellbeing and comfort of the person with dementia. One of the most important team objectives was to involve persons with dementia when possible, and conduct formal and informal ACP conversations with persons with dementia and their family members on a regular basis. All participants reported that a major team action to integrate ACP in daily practice involves defining ACP as a standard agenda point for family meetings, team meetings and one-on-one meetings among HCPs.

### Theme 6. Sharing perspectives & making shared-decisions

Two team leaders, a registered nurse, a quality nurse and a psychologist occasionally noticed the presence of ‘physician dominance’ with respect to sharing perspectives, which limits the opportunity of other HCPs to provide input. Hierarchy was not the only impeding factor, however, as providing input also relies on (mostly nurses) having a proactive and assertive attitude when presenting your voice, and clearly describing situations from the resident’s perspective. Moreover, all team leaders, nurse specialists and a registered nurse also mentioned the lack of competencies regarding PC and ACP held by nurses as regards providing PC for persons with dementia. Correspondingly, physicians and nurse specialists agreed that some nurses indeed have difficulties reporting objectively and tell the story from the resident’s perspective, as they usually offer their own perspective. They felt that some nurses did not feel confident enough to share their perspective, recommendations or advice, and emphasized the importance of including the perspectives of nurses, as they are the closest and core HCPs, and able to actually know the person with dementia. The physicians and nurse specialists explained that they often proactively ask for a nurse’s input so as to hear their voice. Team leaders also explained that in situations where consensus becomes difficult to achieve or there is conflict among the collaborators, they fulfil the role of an independent referee who leads the discussion and aims to seek the best suitable solution.

All participants agreed that reaching consensus starts with providing information, sharing, and understanding diverse perspectives. A transparent atmosphere in which all HCPs have an equal say and a PC culture exists is needed to enable this. The majority pointed out the importance of explaining the consequences of an end-of-life decision to family members. For example, most family members initially did not understand the HCP’s decision to withhold artificial nutrition and hydration during the dying phase. However, when the HCPs explained that doing so would prolong the dying phase and increase the risk of choking and pneumonia, they understood the process of dying better, felt relieved and were able to make appropriate informed shared decisions. Building and cherishing a relationship with the family members is essential in dementia care, because although the participants always wanted to involve the person with dementia in conversations about end-of-life care, this was not always possible due to their cognitive decline.

### Theme 7. From ACP information to actions

All participants discerned the existence of two worlds with respect to transferring information: 1) the participating HCPs at the meeting, and 2) the remaining members of the care team (mostly registered nurses, and certified and uncertified nurses), who need to understand how these agreements were made. In practice, regular formal meetings in which ACP information could be shared include interprofessional team meetings, family meetings, resident meetings, physician-nurse meetings and psychologist-nurse meetings. Resident meetings had different aims and characteristics, all residents were discussed during these meetings from a practical daily life perspective, and most nurses were allowed to participate. In the other meetings, a designated nursing assistant or nurse often participated and represented the group of nurses. Most participants then confirmed that they had short communication lines and explained that they were able to approach each other easily. For example, when participants wish to have a ‘quick chat’ or need ‘quick advice or a decision to be made’, they referred to corridor communications or making phone calls. Some participants suggested that every HCP should take the responsibility to transfer information to their colleagues through verbal communication and documentation. HCPs from one of the care organisations expressed their dissatisfaction at having two different systems: a medical care plan (only accessible to allied healthcare professionals and physicians) and a holistic care plan (only accessible to the nurses). These two systems impeded collaboration, especially with respect to the follow-up of advice, as the nurses were not able to understand the process of decision-making.

### Theme 8. Having a facilitating organisation

The participants distinguished three types of support: team, department and organisation support. Team support involved specialized HCPs such as a PC consultant (HCP specialized in PC) who is able to hold in-depth ACP conversations and provide practical advice on ACP to other HCPs. Participants from one care organisation mentioned that department support could include enabling suitable digital systems, and improving verbal information transfer. On an organisational level, all participants noted that their care organisations consisted of steering groups of diverse HCPs, policy makers or managers, who focus on securing ACP in practice, improving ACP competencies (e.g. arranging e-learning modules about ACP), and supporting the ACP vision.

### Theme 9. Evaluating the IPC process and end-of-life care

Most of the structured evaluation moments referred to decisions taken, tasks performed and care outcomes such as overall end-of-life care for persons with dementia, the satisfaction of family members, and the extent to which HCPs acted according to NH policies. In practice, ad hoc evaluation moments were organised and interprofessional team meetings were utilized to evaluate cases. One team leader explained that their organisation additionally organised quarterly reflection sessions to reflect on structural collaboration and discuss aspects such as task divisions. The participants found it important to ask family members whether they were satisfied with the care given to the person with dementia, and their involvement and role in ACP during regular family meetings. They explained that they conducted family meetings with family members six weeks after the moving day of the person with dementia, and afterwards semi-annually. Some participants even asked family members to participate in interprofessional team meetings for interim evaluations.

#### Reflection meeting

On the basis of the themes above, we discussed a draft version of the guidance document during the reflection meeting (Supplement VIII). We formulated six items based on participant feedback and rephrased these items into six main questions (Fig 3). We show the relationship between the themes of Step Five and the main questions in Supplement IX. Fig 3 provides a summary of the guidance document. The definitive guidance document is provided in Supplement X.

**Fig 3.**
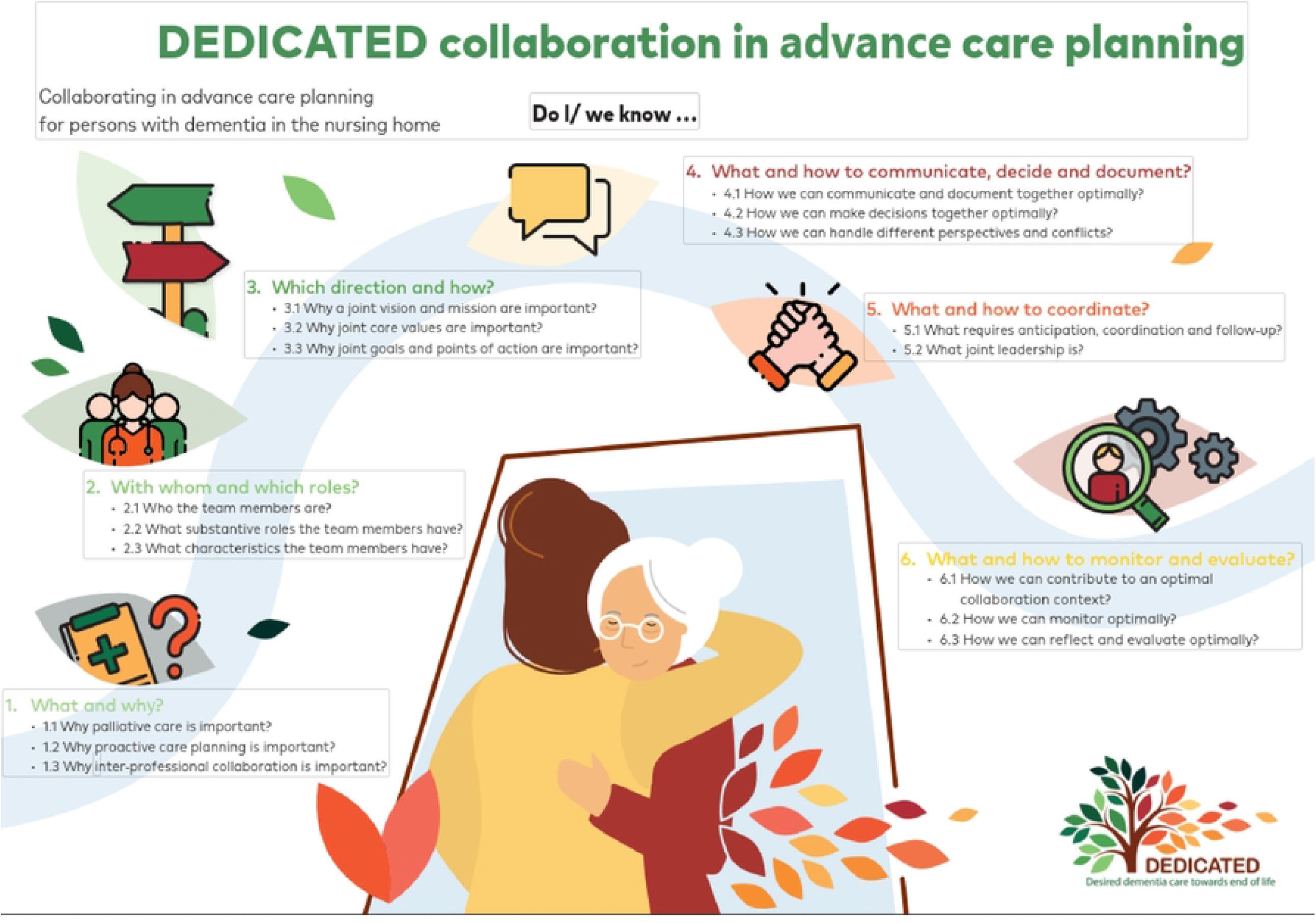
Infographic *DEDICATED-collaborative approach for advance care planning in dementia care*.

## Discussion

Our study showed that nine themes are essential for optimal IPC in ACP for persons with dementia living in NHs. These nine themes resulted in the six elements of the guidance document for IPC in ACP for persons with dementia in NHs. Many studies have emphasized the lack of role clarity among HCPs^9, 60^. This is the first study to outline the similar and complementary roles of diverse HCPs in ACP for persons with dementia. Our finding that nurses should initiate ACP conversations with persons with dementia and their family concurs with Jones et al. (2016)^61^ and Cotter et al. (2018)^62^. This initiating role is best suited to nurses because they often have the opportunity to build a relationship with persons with dementia and their family^63^. This relationship facilitates starting ACP conversations in dementia care^36^ and the PC needs of persons with dementia to be addressed^64^. Moreover, in accordance with Kim et al. (2021), nurses act as care coordinators as they communicate closely with other HCPs to monitor ACP in dementia care^65^. In Dutch NHs, team leaders are seen as following a distinct profession and part of the care team^66^. We demonstrated that team leaders have a leading role in guiding the team vision. Research has shown that creating clarity about the team vision is significantly associated with positive team performance^67^. This could also facilitate role clarification, because developing goals related to the team vision enables each team member to understand the direction in which to move^68^ and to clarify their role in this journey^67^.

An increased common awareness is required among HCPs that dementia is a life-limiting disease and that the integration of a PC approach is necessary in dementia care^69^. We suggest that the awareness of ACP is related to applying a PC approach in dementia care. This is supported by the findings of Beck et al. (2015), who argue it is necessary to apply a PC approach in order to realize the initiation of ACP in dementia care, because they are connected to each other^70^. The literature also indicates improving knowledge of HCPs about ACP as a facilitator for conducting ACP in dementia care^71^. In contrast with Rietze et al. (2015)^72^, our study demonstrated that physicians actively searched for ways to involve nurses in decision-making processes about ACP. This is supported by Kastbom et al. (2019), a study that underlined integrating diverse views in order to receive input and support from each other^73^. Furthermore, as confirmed by van der Steen et al. (2021)^74^, our study showed that physicians wish nurses to have a proactive attitude, be competent and report objectively. Most studies about IPC targeted nurse-physician collaborations^75-77^. In our study, we additionally showed that psychologists share these three collaboration needs. We also discovered that interpersonal aspects such as the confidence^78^ and power differences^75^ described by other studies could also affect IPC in ACP. We did not discuss other factors of influence, however, such as cultural beliefs^79^ or confusion about legal matters^71^.

### Strengths and limitations

This is the first study outlining general and complementary roles of four professions in collaborative ACP in dementia care. We performed investigator triangulation, whereby different researchers and various HCPs provided their input and thereby increased the validity of our analysis^80^. Applying appreciative inquiry also allowed participants to co-create the guidance document in a structured manner^81^. The generalisability of our results is limited, however, because we used a non-probability sampling method^80^ and conducted the research in Dutch NHs. Dutch NHs employ their own staff from various professions, and thus have an intrinsically interprofessional character^82^. Moreover, we did not include other HCPs involved in ACP in dementia care such as NH managers, chaplains and social workers. Nevertheless, this study provides insight into the diverse roles of four different professions in ACP for persons with dementia resulting in a practical guidance document for HCPs.

## Conclusion

Our findings show that the meaning of ACP, timing and continuity of ACP, transferring ACP information prior to the move, creating role clarity in ACP, having a shared vision, sharing perspectives and making shared-decisions, going from ACP information to actions, having a facilitating organisation, and evaluating IPC process and end-of-life care are crucial elements in an interprofessional collaborative approach in dementia care. A major implication of this study is that there is a need to increase the awareness and importance of PC and ACP, build a shared vision and clarify the general and complementary roles of the diverse professions. This study also provided a general and practical description of how to conduct ACP conversations in a collaborative way. We indicated that IPC regarding ACP does not solely concern the NH itself, but also extends to the home care setting. We thus highlighted the need for the optimal information transfer of end-of-life wishes during the move for persons with dementia. Future research should explore the interplay between HCPs from the home care and NH settings concerning ACP in dementia.

## Data Availability

All relevant data are within the manuscript and its Supporting Information files.

